# Towards the elimination of cervical cancer in Tanzania, transdisciplinary science for smarter implementation strategies (TRACCTION): a mixed methods study protocol

**DOI:** 10.1101/2024.07.11.24310271

**Authors:** Grace Mhalu, Mari Dumbaugh, Daniel Kipo, Magreat Somba, Dorcas Mnzava, Julia Bohlius, Sally Mtenga, Sonja Merten

## Abstract

**Introduction:** Cervical cancer is the leading cause of cancer-related mortality for Tanzanian women. Multi-level, intersecting factors prevent access to care along the cervical cancer care cascade. However, there is a paucity of data, especially in non-urban areas, exploring the specificity and nuances of these factors locally, such as perceptions and embodied experiences of cervical cancer, use of traditional medicine, stigma, information generation and circulation and loss to follow up care. TRACCTION is an exploratory, mixed-methods study running from 2023-2025 to expand understanding of barriers to and facilitators of uptake of cervical pre-cancer screening and treatment and cervical cancer care in southeastern Tanzania.

**Methods and analysis:** Research activities include a two-stage randomised cross sectional survey of women 18 years of age and older, qualitative data collection using diverse, community-based methods and a longitudinal public health facility record linkage of patients accessing cervical pre-cancer screening and treatment. By employing a transdisciplinary approach, TRACCTION will inform a participatory process to formulate cervical cancer education and policy recommendations.

**Ethics and Dissemination:** This study received ethical approval in Tanzania and Switzerland. Informed written consent will be obtained from each study participant and a waiver of informed consent was obtained to retrospectively analyse public health facility records related to study objectives. A Policy and Technical Advisory Group, comprised of a diverse group of stakeholders including community members, will co-design a policy and service delivery analysis, followed by recommendations and dissemination plans. Study results for each work package will be published in peer reviewed journals and shared at relevant conferences globally.

## INTRODUCTION

Cervical cancer (CC) is the most commonly diagnosed cancer in Tanzania and the leading cause of cancer-related mortality for Tanzanian women [1]. The underlying cause of CC is infection with high-risk variants of Human Papilloma Virus (HPV) [2]. HIV infection increases the risk for persistent HPV infection, and progression to pre-cancerous cervical lesions and invasive CC [3,4], making women living with HIV six times more likely to develop CC than women who are not living with HIV [2]. In Tanzania, where 1.1 of the 1.7 million adults living with HIV are women, CC is a significant women’s health concern [5].

A regional demonstration program to vaccinate girls against HPV was rolled out in Tanzania from 2014-2016, and in 2018 the HPV vaccine was integrated into the country’s national immunization schedule [6]. However, modelling studies show that it will take decades until HPV vaccinations will effectively reduce CC incidence and mortality; therefore, prioritizing screening for and treatment of cervical pre-cancer remains paramount to addressing this public health concern [7]. The *Tanzanian Service Delivery Guidelines for Cervical Cancer Prevention and Control Programme* stipulate cervical pre-cancer screening, HPV testing and follow-up treatment protocols for women aged 30-49 and women aged 15-49 living with HIV [8].

Existing evidence from Tanzania identifies structural, systemic, socioeconomic and sociocultural barriers to the uptake of cervical pre-cancer screening [9], resulting in a national screening coverage in 2018 of only 11% of women aged 30-50 [10]. We have identified gaps in the evidence base which, if explored further, could help inform ongoing efforts to increase uptake of cervical pre-cancer screening and detect and treat cervical pre-cancer and CC at earlier stages. First, most CC evidence from Tanzania is from urban settings where health services generally, and cancer services in particular, are more prevalent and accessible [9]. Many existing studies also focus exclusively on women living with HIV [9]. While women living in urban settings and women living with HIV are indeed priority populations, higher rates of CC are recorded in rural areas in different regions globally [11] and CC is a relevant concern to women living with and without HIV. Therefore, more insight into cervical pre-cancer and CC knowledge and practices of women living with and without HIV in non-urban settings is needed to develop and refine policies relevant to different populations. In addition, in many Tanzanian health facilities routine attendance to cervical pre-cancer screening and follow up services is documented using paper-based registries. Though inferences drawn from such programmatic data can contribute to the effective monitoring and management of CC services, paper-based documentation and data are not always easily accessible to effectively monitor and manage service provision and longitudinal patient follow up care. More systematic data collection systems, such as those which can link and track patient contacts with and loss to follow up from the health system over time, could contribute to improvements in service outreach, access and patient retention [12].

Understanding perceptions and embodied experiences of disease within specific sociocultural contexts is also known to provide insight into health behaviour [13–15], including knowledge and sense making related to cervical cancer [16,17]. A number of intersecting factors, “a constellation of socio-medical considerations” [17], contribute to embodied experiences related to health and health behaviour [14]. There is, however, a paucity of data exploring these themes in relation to CC in Tanzania. For example, given high rates of traditional and alternative medicine use in Tanzania [18], insight into diverse pathways of CC prevention and care, including the use of home-based care, traditional healers or religious and prayer healing, is lacking. Stigma also plays an important role in cervical pre-cancer screening avoidance in other similar contexts [19,20], but literature on the intersection of conceptions of CC, pre-cancer screening and stigma in Tanzania is sparse [9]. Finally, social networks are shown to facilitate information generation and dissemination [21,22], but our knowledge of how these dynamics play out related to CC, especially in low-income settings such as Tanzania, is limited [23,24].

*Towards the elimination of cervical cancer in Tanzania: Transdisciplinary science for smarter implementation strategies* (TRACCTION) is a mixed methods exploratory study running from 2023-2025 to expand the depth and breadth of the understanding of multi-dimensional, intersecting barriers and facilitators of uptake of cervical pre-cancer screening and treatment and CC care in non-urban settings in southeastern Tanzania. By employing a transdisciplinary approach [25], this study will inform a participatory process involving a variety of stakeholders to ultimately formulate CC education and policy recommendations.

## METHODS AND ANALYSIS

Data collection for the TRACCTION study will take place in communities and public health facilities in a town and surrounding rural areas in Kilombero district, Morogoro region, Tanzania. The study sites are located in southeastern Tanzania in the Kilombero River Valley, where agriculture is the main economic activity. We will collect data in a small but rapidly growing town which, in 2012, had a population of over 106,000 people [26], a village on the periphery of the town and a remote rural village in the same region. The multi-country, interdisciplinary study team comprises local and international health social scientists, clinicians, epidemiologists, and health systems and policy experts to address the first delay of CC screening uptake, construct and analyse a CC prevention and care cascade, and investigate the health and psychosocial needs of women living with CC.

The study is organised into four inter-related work packages (see **Table 1**). Each work package will be conducted successively with results from early research activities iteratively informing the design and implementation of subsequent research activities. We will engage community members, including women of reproductive age, and health system stakeholders from both biomedical and traditional perspectives in transdisciplinary research. This process will culminate in the development of robust education and implementation strategies and policies that aim to eliminate CC in Tanzania.

**Table 1.** Objective, methodology, data source and analysis of each TRACCTION sub-study.

|  | Objective | Methodology | Sample population/<br>data source | Analysis |
| --- | --- | --- | --- | --- |
| **Sub-study 1:**<br>**Qualitative**<br>**exploration**<br>**and cross**<br>**sectional**<br>**community**<br>**survey** | To identify demand side gaps and opportunities for increasing cervical pre-cancer screening uptake. | *Convergent mixed-methods*<br>Focus group discussions; in-depth interviews; non-participant observations; social network analysis<br>Two-stage randomised quantitative survey | <u>Qualitative</u><br>Purposive sampling of key stakeholders:<br>*Focus group discussions*: women in general population; women living with HIV; women who have attended screening; women who have not attended screening<br>*Key informant interviews*: Healthcare providers from facilities with and without cervical pre-cancer screening<br>*In-depth interviews*<br>Women from the general population; women and men identified through social network analysis.<br>*Observations*: Facility- and community-based cervical pre-cancer screening clinics<br>*Social network analysis*: Purposively selected women of different levels of education who have and have not access cervical pre-cancer screening services | <u>Qualitative</u><br>Inductive, iterative, constant comparative approach (constructivist grounded theory)<br><u>Quantitative</u><br>Descriptive statistics.<br>Multi-level, uni- and multivariate logistic regression to estimate associations of predictors with screening and CC knowledge.<br>Latent class analysis to test theoretical assumptions about the relevance of information networks, and associations of classes with uptake of screening. |
|  |  |  | *Quantitative*<br>Stratified, two-stage randomised cluster sample of women using probabilistic sampling at three sites (secondary town, periphery village, remote village) |  |
| **Sub-study 2: CC screening record linkage** | To document the CC care cascade from cervical pre-cancer screening through treatment for women living with and without HIV. | Longitudinal record linkage for patients accessing cervical pre-cancer screening and treatment. | Paper-based patient records from facility- and community-based cervical pre-cancer screening clinics at a government health center (2011-2023). | Descriptive statistics and cumulative incidence rates at each stage of the CC prevention and care cascades.<br>Uni- and multivariable Cox regression analyses to calculate cause-specific hazard ratios for factors associated with reaching cascade stages and developing CC. |
| **Sub-study 3: Patient perspective of CC care through body mapping** | To explore the patient perspective of CC care. | Key informant interviews with district-level health managers and health providers from public and private health facilities.<br>Observations in public and private health facilities.<br>Multi-staged, participatory, visual art-based qualitative methodology. | Purposive sampling of health managers and providers from public and private facilities.<br>Purposive sampling of women diagnosed with cervical pre-cancer and CC. | Inductive, iterative, constant comparative approach (constructivist grounded theory) [27]. |
| **Sub-study 4: Policy and service delivery analysis and strategy development** | To identify policy-related factors that can be improved to promote CC screening and prevention behaviour in the context of universal healthcare. | Policy and program document review.<br>Policy and practice guidance workshops. | Policy and program documents from East and Southern Africa.<br>Women from urban and rural study sites; women living with HIV; men as partners/spouses of women; community leaders including traditional practitioners and religious leaders; government representatives.<br>National level experts, policymakers and clinical academicians. | Content analysis of policy and program documents.<br>Content analysis of detailed field notes from participatory workshops with multi-level stakeholders. |

Each work package is summarised in **Table 1**. and briefly described below.

### Patient and public involvement

By using a transdisciplinary approach, our study aims to centre patient voices and experiences throughout research activities. Patients are first involved in the study during formative qualitative research activities. During these activities, feedback from patients and members of the target population will be used to further develop and refine the approaches and specific topics covered in quantitative and qualitative data collection and analyses. Community health workers, who are members of the communities in which research will take place and are well-informed of community health priorities and concerns, will assist with recruitment for community-based research activities. Finally, a key part of this research is the convening of stakeholders (see Work Package 4 below), including patients and other community members, to give feedback on the research methods, discuss and disseminate study outcomes, and develop relevant educational and policy actions as next steps.

### Sub-study 1: Qualitative exploration and community household survey

We will collect formative qualitative data for a baseline understanding of cervical pre-cancer screening and treatment services available in the study area. In-depth interviews and focus group discussions with women (n=100) and men (n=30) of reproductive age from a variety of socioeconomic, religious and educational backgrounds will explore CC knowledge, patterns of resort and barriers and facilitators to cervical pre-cancer screening and CC care. Qualitative research activities will also delve into local, embodied experiences related to CC: conceptualisations and perceptions of female anatomy, CC disease pathology, cervical pre-cancer screening procedures and CC-related stigma, including HIV-related stigma.

To better understand how cervical pre-cancer services are experienced by women and the health workers delivering them, we will conduct observations of screening services at public and private health facilities and during community outreach events. We will also complete key informant interviews (KIIs) with health workers and health facility administrators (n=12) involved in cervical pre-cancer screening and treatment services as well as HIV services. KIIs will occur in the language preferred by the participant (Kiswahili or English). In addition, we will conduct KIIs with traditional practitioners across study sites (n=12). Qualitative sample sizes have been determined from relevant literature [28] and previous experience to reach data saturation; if saturation is not reached once anticipated sample sizes are met, we have the option to conduct additional qualitative data collection as necessary. Audio recordings of all interviews and focus group discussions will be translated and transcribed verbatim into English. During observations the researcher will take discreet notes, and fully elaborate notes in English after the observation is complete.

We will use an iterative, constant comparative approach (constructivist grounded theory) [27] for the analysis of all qualitative data including transcriptions and observation notes. Triangulating information from women, men and community- and facility-based healthcare providers, we will develop a theoretical framework for cervical pre-cancer screening uptake in Kilombero district, revisiting and applying a gender analysis to the conceptualization of access to healthcare [29].

A two-stage randomised community survey will be developed from a review of cervical pre-cancer screening literature and previously used surveys from Tanzania and other east and southern African contexts [30–33]. We will use initial results from formative qualitative activities described above to adapt survey questions and translations into Kiswahili to local beliefs, perceptions, epistemic conceptualisations and health service organisation. Topics covered in the survey are detailed in **Table 2**. Survey questions will be translated into Kiswahili by the research team and local research assistants during translation workshops.

**Table 2.** Topics covered in TRACCTION community survey.

|  |  |
| --- | --- |
| • Sources of and access to health information, including cervical cancer<br>• Cervical cancer and cervical pre-cancer screening knowledge and perceptions<br>• Access, affordability and use of health services including cervical pre-cancer screening and CC treatment<br>• Quality of health services | • Cervical cancer- and HIV-related stigma<br>• Self-efficacy<br>• Social support and couple cohesion<br>• Socio-demographics including household assets and wealth<br>• Household decision making<br>• Insurance coverage |

Experienced, local research assistants will collect survey data. Before data collection begins, the team will participate in a week of survey training and pretesting, including refresher training on research ethics, confidentiality and informed consent. A pilot study will be conducted in the study region in a village an adequate distance from study sites to avoid study contamination. After the pretest and pilot studies, the study team and research assistants will work together to refine the wording, translations and length of the questionnaire.

Through probabilistic sampling using a stratified two-stage cluster design [34], we will select households at three research sites: a town, a periphery village and a remote village in Morogoro Region, Tanzania. We will administer the survey to one consenting woman aged 18 and over per household in each of the randomly selected clusters. Research assistants will confirm survey eligibility and obtain informed written consent from each survey participant before the survey begins. Data will be collected using Open Data Kit software on password-protected electronic tablets. Each survey participant will be identified on the server with an anonymized participant ID to maintain participant anonymity and confidentiality. Each evening after field work concludes survey data will be sent to a secure server in Tanzania and Switzerland via mobile network. Only select members of the core study team will have access to participant data.

To analyse survey data we will calculate frequencies and prevalence of CC screening and service use, followed by multilevel, univariate and multivariable logistic regression analyses to estimate associations of selected predictors with cervical pre-cancer screening in the past 12 months (main outcome variable). Secondary outcome variables include any cervical pre-cancer screening and knowledge of CC symptoms. Main predictors will include the decision to use screening services, delays in reaching and, ultimately, receiving the service. Other survey outputs include the clarification of information sources for CC (social network and information outcomes), barriers and facilitators of cervical pre-cancer screening uptake and opportunities for strengthening cervical pre-cancer service delivery. We will also test hypotheses generated by qualitative analysis, and anticipate using latent class analysis to test theoretical assumptions about the relevance of information networks [35]. Latent classes of information groups will be created based on variables capturing the importance of information sources. Associations of these classes with uptake of cancer screening will be investigated with STATA MP (College Station, TX: StataCorp LP) and R (R Foundation for Statistical Computing, Vienna, Austria).

Initial analysis of the quantitative survey, especially data on sources of health information, will inform our approach to a qualitative social network analysis [21,22,36]. The social network analysis will allow us to explore how information on perceptions and knowledge of CC, cervical pre-cancer screening and CC services is diffused, received and interpreted by women, their partners and their closest social networks in the community. We will also identify opinion leaders and decision makers related to women’s health and CC within women’s social networks in different socio-geographic contexts (e.g. town versus rural communities).

We will begin the social network analysis by selecting six women, or ‘egos’, who participated in formative qualitative activities in each of the three research sites. Women will be purposively selected for diverse educational backgrounds and will be asked to list all sources of CC information including people, institutions (e.g. health facility, church, community board), media sources (e.g. radio, social media, etc.), and other places (e.g. women’s groups, markets, hairdressers, restaurants/bars). We will then ask participants to rank their information sources in order of importance, and the top five information sources for each initial participant (n=30) will be interviewed about their own health-related social and information networks and sources, as well as knowledge on CC and cervical pre-cancer screening and uptake (women only).

We will qualitatively analyse the social network analysis data, graphically displaying the social networks of the primary ‘egos’ [21]. To elucidate the reasoning of whether, why and where women make use of cervical pre-cancer screening, we will identify how information about cervical cancer is circulating between sources (people, institutions, media and other sources). This will help us to better understand how different groups of people of similar and varying positionalities obtain and interpret information about CC. The intersections and interactions between biomedical and traditional (e.g. spiritual, religious, herbal and traditional medicine) health knowledge and systems will especially be considered. We aim to create a typology of information networks or ‘epistemic communities’ [37,38], which we will re-examine and validate with the quantitative analysis.

### Sub-study 2: Health facility record linkages

Data from patient records will be extracted from existing, paper-based health facility registers and linked longitudinally to document the CC care cascade from cervical pre-cancer screening through treatment for women living with and without HIV. We will conduct this data collection and analysis in a public, government-run secondary health facility with patient records from 2014-2023. Linkage variables will include surname, first name, date of birth or age, home village and where available numeric patient identifier. Other linkage variables will be explored. Exposure and outcome variables for cervical pre-cancer screening will include among others: age at screening visit, type of service (first time screening; follow-up visit; re-screening) and date of visit, visual inspection with acetic acid screening and result, human papillomavirus screening and result, treatment, biopsies and results thereof, referral and calendar year.

To link patient records, we will use patient identifying information (see linkage variables above) to identify records belonging to the same person, using deterministic and probabilistic record linkage methods [39]. The record linkage process will include: a) *preprocessing*, b) *deterministic matching*, c) *blocking*, d) *matching*, e) *post-processing* and f) *validation* [39]. All records belonging to a unique person will be assigned a project-specific, unique linkage ID.

In the first stage of the linkage process, *pre-processing*, we will harmonise linkage variables in terms of format, structure, and content using regular expression templates. Dates (e.g. date of birth) will be checked and put into a standard format. For name standardisation, we will benefit from experiences from our previous record linkages in South Africa and Malawi [40–44]. We will use *deterministic deduplication* (also known as *exact matching*) to link records that have identical key variables of first name, surname, and date of birth [39]. *Blocking* is the systematic restriction of comparisons to ‘blocks’ of plausible pairs, where one or more variables match [39]. We will apply *blocking* strategies to identify potential pairs which will increase the efficiency of the linkage process.

At the *matching* stage, we will calculate match (m-probability) and nonmatch (u-probability) for each outcome. The likelihood of a match for each variable is transformed into a weight and all weights are summed up to a total outcome weight for each pair of records [39]. We will examine the range of the total weights to determine upper and lower cut-off thresholds and reject pairs with a total weight below the lower threshold [39]. During *post-processing,* we will clerically review possible pairs and make a determination on the final match status [39]. To increase reliability at this stage, two reviewers will review the possible pairs. Finally, we will *validate the linkages*. Given we do not have unique patient identifiers within or across existing databases, we will validate our linkages with external information such as the proportion of women living with HIV having received cervical pre-cancer screening, the proportion of positive Visual Inspections with Acetic Acid, the proportion of women referred for further services and followed-up.

If systematic data gaps are observed we will discuss potential reasons for these gaps with staff at the cervical pre-cancer screening unit, and work together to create systems which may prevent these gaps in future. If we cannot obtain additional data we will compare the characteristics of linkage variables that are missing to those that are not missing to detect potential biases. If potential biases are identified we will describe these and conduct sensitivity analyses to judge the impact on our calculations. For missing analyses variables we will impute missing data assuming these occurrences at random.

After the linkages are complete, we will retain the linkage IDs and remove all patient identifying information (e.g. patient IDs and names). The product of the record linkages will be a CC screening cohort. The newly created cohort databases will be used to document two CC-related cascades of care for women living with and without HIV accessing services in a government-run secondary health facility in Morogoro Region, Tanzania: 1) Cervical pre-cancer screening cascade and 2) Preventative treatment cascade. All cascade stages will be aligned with the Tanzanian CC control policies [10]. All HIV-positive and -negative women aged ≥18 years will be included in analyses to determine the number of women at each of the linked stages (see **Table 3**.).

**Table 3.** Linked stages included in Cervical Cancer Screening and Preventative Treatment Cascades [45].

| **Screening Cascade** |  |
| --- | --- |
| I. | Screened for cervical pre-cancer |
| II. | Documented negative result at first screening |
| III. | Re-screened |
| IV. | Documented negative screening result at re-screening |
| **Preventative Treatment Cascade** |  |
| I. | Documented positive result at first screening |
| II. | Referred for diagnosis |
| III. | Linked to treatment |
| IV. | Retained and re-screened |
| V. | Documented negative result at re-screening |

We will calculate descriptive statistics to describe baseline patient characteristics. For all stages of the cascade we will calculate cumulative incidence and percentages with 95% confidence intervals. We will use uni- and multivariable Cox proportional hazard regression analyses to calculate cause-specific hazard ratios with 95% CIs for factors associated with reaching cascade stages. We will use STATA MP (College Station, TX: StataCorp LP) and R (R Foundation for Statistical Computing, Vienna, Austria) for all analyses.

### Sub-study 3: Addressing treatment delays to ensure continuum of care for CC

To investigate women’s therapeutic journeys, including delays and loss to follow-up as well as the health and psychosocial needs of women with cervical pre-cancer and cancer, we will engage in unstructured observations at four healthcare facilities at different levels in the district, focusing our observations on CC-relevant routines and practices. We will purposively select patients (N=10) attending services at a CC clinic who have been diagnosed with cervical pre-cancer or CC stage 1 or higher to follow, with their consent, their patient experience at the facility on that day. Researchers completing observations will take discreet notes throughout the observation, and fully elaborate notes in English after the observation.

Then, we will conduct KIIs with Kilombero district-level health managers and qualified healthcare providers from the public and private sector (N=8) who are involved in specialised CC treatment. KIIs will focus on health care managers’ and providers’ knowledge and perceptions of CC including CC prevention, the availability and acceptability of critical inputs (e.g. human resources, supplies, equipment, infrastructure, finances) as well as referral procedures and appointment-setting. Interviews will be held in Kiswahili or English at the direction of the participant.

Informed by data gathered during interviews and facility observations, we will then facilitate two series of structured group discussions with women living with CC (n=6-8 participants per series) using the art-based research method of body mapping [46,47]. Participants will engage in five sessions of art- and discussion-based exploration and exchange. In each facilitated session, participants will develop a new theme linked to CC including their personal embodied experiences with CC, their therapeutic journey, the impact CC has on their lives and their strategies for self-care and coping. Discussions during the sessions will be held in Kiswahili and audio recorded with participants’ permission. All audio recordings will be translated into English and transcribed verbatim and, along with visuals produced during the sessions, written notes and observational data, will be retained for analysis.

We will employ constructivist grounded theory [48] for the analysis of the data collected during the health system stakeholder KIIs, CC services observations, and group body mapping sessions. Combined with KIIs and observations, individual visual and narrative data from body mapping will allow us to establish detailed patient journeys through CC services at both public and private facilities. Each strand of qualitative data will be separately analysed, then juxtaposed and triangulated against each other. Finally, we will hold a workshop to discuss and validate analyses and findings from across different qualitative data collection methods with representatives of the study participants, including one representative from each body mapping group and representatives of health professionals included in research activities. Ultimately, qualitative work will identify gaps and opportunities for educating and linking women to cervical pre-cancer testing, follow up, treatment and CC care.

### Sub-study 4: Identifying additional policy factors that can be improved to promote CC screening and prevention behaviours

In a final stage of our study, we will undertake a policy and service delivery co-design process to identify context-specific opportunities for the provision of cervical pre-cancer screening and therapy, to align or (re-)design health service delivery approaches that address gaps in the continuum of care, to define policies promoting equitable access to CC-related services such as HPV vaccination, screening, and CC treatment, and to create linkages to existing universal health coverage efforts in Tanzania, ensuring CC prevention and treatment are included. The overall work package will be supported by the Policy and Technical Advisory Group (PTAG) which will consist of national-level experts, policymakers, clinical academicians and research participants, and research team members. The PTAG to co-design a policy and service delivery analysis followed by recommendations and a dissemination plan for each stakeholder group for uptake by national government stakeholders and policymakers.

A policy analysis of CC in the east and southern Africa regions will inform the policy and intervention strategy analysis. We will conduct a political economy analysis [49–51] to assess opportunities and challenges to improving the enabling political environment for CC prevention and treatment, and inclusion of CC within efforts to establish universal healthcare in Tanzania. Based on the findings of the regional and national policy and practice analyses, we will support the PTAG to convene a participatory stakeholder meeting where we will present the results of the policy analyses. In this workshop, the policy environment will be critically assessed, stakeholders will discuss re-adaptations of the design of vaccination campaigns and cervical pre-cancer screening programs, and develop new ideas to enable equitable access to information and uptake of CC prevention and therapy. Stakeholder participants in the policy guidance workshops will include women from all study sites (purposively selected for geographic, socio-economic and educational diversity), women living with and without HIV, men as partners/spouses of women, community leaders (including traditional practitioners and religious leaders), health workers including community health workers and government representatives.

## ETHICS AND DISSEMINATION

We have received approval for this study from ethics committees in both Tanzania (Internal Review Board of the Ifakara Health Institute and the National Institute for Medical Research) and Switzerland (Ethikkommission Nordwest-und Zentralschweiz/EKNZ). Before data collection begins for each research activity, data collectors will explain, in detail, the aims and parameters of the TRACCTION study in either Kiswahili or English, whichever language is preferred by the participant. Data collectors will then give participants an information sheet detailing the same information explained to them and obtain informed written consent to participate in the study. For qualitative research activities (Work Packages 1 and 3), participants will also be asked to give verbal consent to be audio recorded. If qualitative participants do not wish to be audio recorded, researchers will continue with research activities as planned but take detailed notes of the exchange by hand. The only exception to obtaining informed written consent from participants is for Work Package 2, wherein we expect to analyse up to approximately 17,000 patient records collected since 2014. Given that retrospectively obtaining informed consent to use a dataset of this size is not feasible from paper-based systems with a lack of patient contact information, we have obtained ethical approval in both Tanzania and Switzerland for a waiver of informed consent to retrospectively analyse specific health facility records related to the TRACCTION study objectives (further use of data). For Work Package 4, we will provide each PTAG participant with an information sheet outlining the objectives of the participatory workshops and the roles envisioned for stakeholders. We will obtain verbal informed consent from all individuals invited to participate before beginning workshop discussions.

Prior to the commencement of the study, kick-off meetings with various community, health facility and government stakeholders will be held to ensure that stakeholders are aware of the project and main project objectives. Our dissemination plan includes various modalities tailored to each stakeholder group to ensure that key messages generated from our study results are accessible to local communities, patients, health care professionals and other stakeholders. For example, PTAG workshops aim to produce systems-level policies and strategies informed by study results while also developing materials on CC information and related services and strategies for distribution which respond to the stated needs of the communities. In addition, peer-reviewed articles related to results from each work package will be written to ensure that study results are disseminated to the wider scientific community and can inform future CC-related policy and program development in similar contexts.

This study will offer much needed insight into women’s perceptions, embodied experiences with and access to cervical pre-cancer screening and CC diagnosis, treatment and care services as well as lay groundwork for future research in the field. Lessons learned from this study could be applied to research in different geographic and/ or health facility contexts, thereby contributing to targets of Sustainable Development Goal 3 which aim to reduce premature deaths from non-communicable diseases and ensure sexual and reproductive health care services are universally accessible globally.

## Data Availability

All data produced in the present study are available upon reasonable request to the authors

## Acknowledgments

The authors would like to thank our administrative colleagues at the Swiss Tropical and Public Health Institute and the Ifakara Health Institute for all of their efforts which facilitated the funding and launch of this study. Thanks also to Professor Dr. Maja Weisser Rohacek for her contributions to the original study protocol and kindly facilitating the study team’s introduction to the local research community.

## Author statement

SM conceived of and developed the original study protocol, with JB leading the development of Work Package 2. SM and DN made significant contributions to the finalisation of the protocol, including finalising the study sites. GM and MD wrote the protocol manuscript for publication. DK and MS edited and finalised this manuscript. All authors contributed to the finalization of the study protocol.

